# Rapid detection of SARS-CoV-2 B.1.1.529 (Omicron BA.1) variant by SYBR Green□based RT□qPCR

**DOI:** 10.1101/2023.05.16.23289717

**Authors:** Fadi Abdel-Sater, Rawan Makki, Alia Khalil, Nader Hussein, Nada Borghol, Ziad Abi Khattar, Aline Hamade, Nathalie Khreich, Mahoumd El Homsi, Hussein Kanaan, Line Raad, Najwa Skafi, Fatima Al-Nemer, Zeinab Ghandour, Nabil El-Zein, Mohamad Abou-Hamdan, Haidar Akl, Eva Hamade, Bassam Badran, Kassem Hamze

**Affiliations:** Laboratory of Molecular Biology and Cancer Immunology (Covid 19 Unit), Faculty of science I, Lebanese University, Hadath, Beirut, LB 1003

**Author notes:** Co-corresponding authors **Corresponding author:** Kassem Hamze: Laboratory of Molecular Biology and Cancer Immunology (Covid 19 Unit), Faculty of Science I, Lebanese University, Hadath, Beirut, LB 1003. Bassam Badran: Laboratory of Molecular Biology and Cancer Immunology (Covid 19 Unit), Faculty of Science I, Lebanese University, Hadath, Beirut, LB 1003.

**Keywords:** Omicron variant, COVID-19 pandemic, SARS CoV-2, SYBR Green-based RT-qPCR

## Abstract

The COVID-19 pandemic is unceasingly spreading across the globe, and recently a highly transmissible Omicron SARS-CoV-2 variant (B.1.1.529) has been discovered in South Africa and Botswana. Rapid identification of this variant is essential for pandemic assessment and containment. However, variant identification is mainly being performed using expensive and time-consuming genomic sequencing. In this study we propose an alternative RT-qPCR approach for the detection of the Omicron BA.1 variant using a low-cost and rapid SYBR Green method. We have designed specific primers to confirm the deletion mutations in the spike (S Δ143-145) and the nucleocapsid (N Δ31-33) which are characteristics of this variant. For the evaluation, we used 120 clinical samples from patients with PCR-confirmed SARS-CoV-2 infections, and displaying an S-gene target failure (SGTF) when using TaqPath COVID-19 kit (Thermo Fisher Scientific, Waltham, USA) that included the ORF1ab, S, and N gene targets. Our results showed that all the 120 samples harbored S Δ143-145 and N Δ31-33, which was further confirmed by Whole genome sequencing (WGS) of 10 samples thereby validating our SYBR Green-based protocol. This protocol can be easily implemented to rapidly confirm the diagnosis of the Omicron BA.1 variant in COVID-19 patients and prevent its spread among populations, especially in countries with high prevalence of SGTF profile.

## 1. Introduction

The World Health Organization has declared the COVID-19 outbreak in China a global pandemic on March 2020. Since then, the causative agent, severe acute respiratory syndrome coronavirus 2 (SARS-CoV-2), has been extensively investigated, with more than 8,313,147 million genomic sequences obtained and made freely available on the GISAID database on February 16, 2022. As the SARS-CoV-2 spreads over the globe, a natural process of random mutations and evolution continues (Habas et al., 2020, Li et al., 2020). In some circumstances, a mutation provides an evolutionary advantage for the virus, resulting in the emergence of a novel viral lineage that outnumbers previous forms. Many SARS-CoV-2 variants have emerged I various regions of the world, and five of them, Alpha (B.1.1.7), Beta (B.1.351), Gamma (P.1), Delta (B.1.617.2), and, most recently, Omicron (BA.1), have been designated as variants of concern (VOC) due to their increased transmissibility, virulence, and/or ability to evade immunity (Davies et al., 2021, Faria et al., 2021, Tagally et al., 2021, Otto et al., 2021, Cele e al., 2022, Planas et al., 2022). Because of its unusual mutational profile and rapid increase in prevalence, the Omicron variant, first found in South Africa and Botswana in November 2021, acquired its VOC classification within days, displacing pre-existing lineages in that country. Omicron has since surpassed pre-existing variants in Europe, the United States, and a number of other countries, causing a new worldwide outbreak. Omicron (BA.1) displays more than 55 mutations in its genome, with 33 of them occurring only in the spike(S) protein among which three deletions (Δ69-70, Δ143-145, Δ211) and one insertion (ins214EPE). Also, the nucleocapsid (N) protein contains four mutations including one deletion Δ31-33, and three substitutions (P13L, R203K, G204R) (Planas et al., 2022, Callaway, 2021).

The rapid spreading of Omicron across the globe urge the need to establish a rapid and low-cost diagnostic tool that could quickly and efficiently detect and track it in order to initiate response and proper policy for pandemic containment. Until now, sequencing is considered as the gold-standard method for identifying SARS-CoV-2 variants. This method is accurate but time-consuming and expensive. Alternatively, rapid and low-cost RT-qPCR detection methods were proposed recently including the use of a range of primers specific for mutations common to VOCs (Abdel Sater et al., 2021, Corman et al., 2020, Borsova et al., 2021, Granato et al., 2021, Zelyas et al., 2021, Vega-Magana et al., 2021, Bedotto et al., 2021), or utilizing commercial kits such as TaqPath COVID-19 diagnostic tests (Thermo Fisher Scientific, Waltham, USA) that included the ORF1ab, S, and N gene targets. Some of the SARS-CoV-2 VOCs, Alpha (B.1.1.7) and Omicron (BA.1), generate dropout of the S-gene result in TaqPath kit, with positive results for the other targets (ORF1ab and N genes). This feature has been used as an indicator or screening method to identify these particular variants. The failure of the S-gene target is caused by a deletion mutation Δ69-70 in the respective gene and is called the S-gene target failure (SGTF). Following the use of TaqPath kit, confirmation of the Omicron variant can be performed by specific RT-PCR assays targeting mutations that are characteristic to this variant (S Δ143-145 and N Δ31-33) with mutation-matched primers or probes, and identification through amplification and melting curve analyses (Dachert et al., 2021). However, at least a subset of samples should be further characterized by sequencing to increase the confidence and reliability of the obtained results [15].

In 16 Febuary 2022, 1,200,309 Omicron sequences have been shared worldwide through the GISAID, among them 1,129,379 sequences belong to the BA.1 sub-variant (94,09%), and all harboring the S Δ143-145 and N Δ31-33 mutations. Consequently, we report here a low-cost SYBR Green-based qPCR protocol for the rapid and specific detection of these two deletion mutations. We propose primer sets, specific to S Δ143-145, and N Δ31-33, that could be applied as a second step to confirm the diagnosis when the number and proportion of SGTF are steadily increasing when using the Applied Biosystems TaqPath RT-PCR COVID-19 kit (Thermo Fisher Scientific, Waltham, USA).

## 2. MATERIALS AND METHODS

### 2.1. Clinical specimens

220 clinical nasopharyngeal samples were collected and selected for this study from 03 to 22 December. All these samples were previously tested for the presence of SARS-CoV-2 at the Laboratory of Molecular Biology and Cancer Immunology of the Lebanese University, using the Applied Biosystems^™^ TaqPath^™^ COVID-19 assay which targeted the RdRp, N and S genes. 120 of these clinical samples were positive for SARS-CoV-2 by TaqPath kit with SGTF profile and 100 clinical samples were negative for SARS-CoV-2. Written informed consent was provided by all participants.

### 2.2. RNA extraction and SARS-CoV-2 qRT-PCR

RNA was extracted from 200 μL of VTM from the clinical samples on Kingfisher flex purification system (Thermo Fisher) using MagMAX^™^ Viral/Pathogen Nucleic Acid Isolation Kit (Thermo fisher). Reactions were performed in a 20 μL final volume reaction containing 5 μL of extracted RNA. RT-qPCR was performed using QuantStudio 5 real-time PCR detection system (Thermo fisher) and TaqPath 2019-nCoV real-time PCR kit (Thermo fisher), which targeted the RdRp, N and S genes of SARS-CoV-2.

For the validation of Omicron variant in SARS-CoV-2 positive samples, total RNA was retrotranscribed into cDNA using iScript cDNA synthesis kit (BioRad), following the manufacturer’s recommended procedures. Quantitative RT-PCR was carried out using iTaq universal SYBR Green super mix (Bio Rad). Real-time PCR was performed using Bio Rad CFX96 Real□Time PCR Machine. The thermal cycling conditions used were as follows: 94 °C for 2 min, followed by 40 cycles of amplification at 94°C for 10 seconds, and 60°C for 1 minute. The reaction was completed by determining the dissociation curve of all amplicons.

### 2.3. Primer design

Primers were designed based on the full sequence of the Wuhan SARS-CoV-2 genome sequence from NCBI nucleotide database (<u>NC_045512</u>) and the full sequence of B.1.1.529 (BA.1) variant (EPI_ISL_6640917) from GISAID EpiCoV database (Fig 1). Bioinformatics tools were used to design and verify the SARS-CoV-2 specific primers. We validated our PCR primers *in-silico* with the PCR Primer-Blast tool which allows to investigate the amplification targets of primers to thereby ensuring adequate specificity. The primer sets used in this study were synthesized and delivered by Macrogen (Republic of Korea) (Table 1).

**Table 1:**
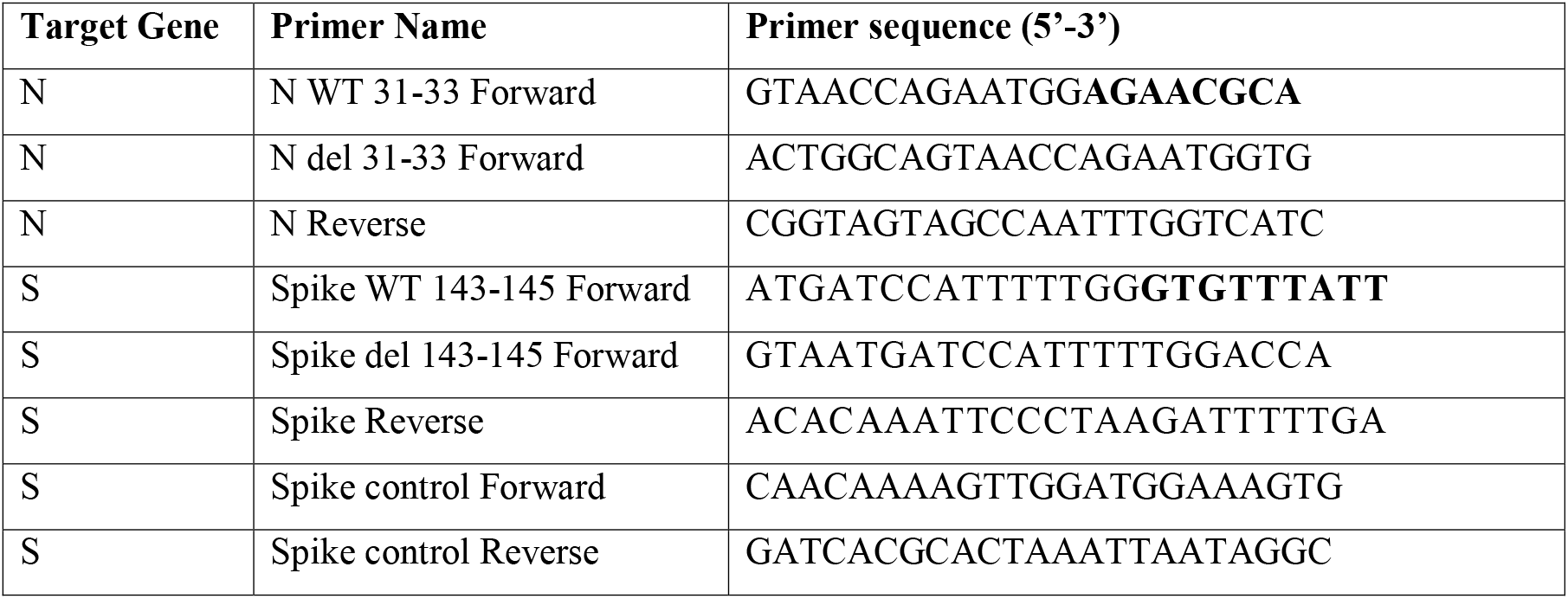
List of primer sets. Deleted nucleotides are in bold.

**Fig 1.**
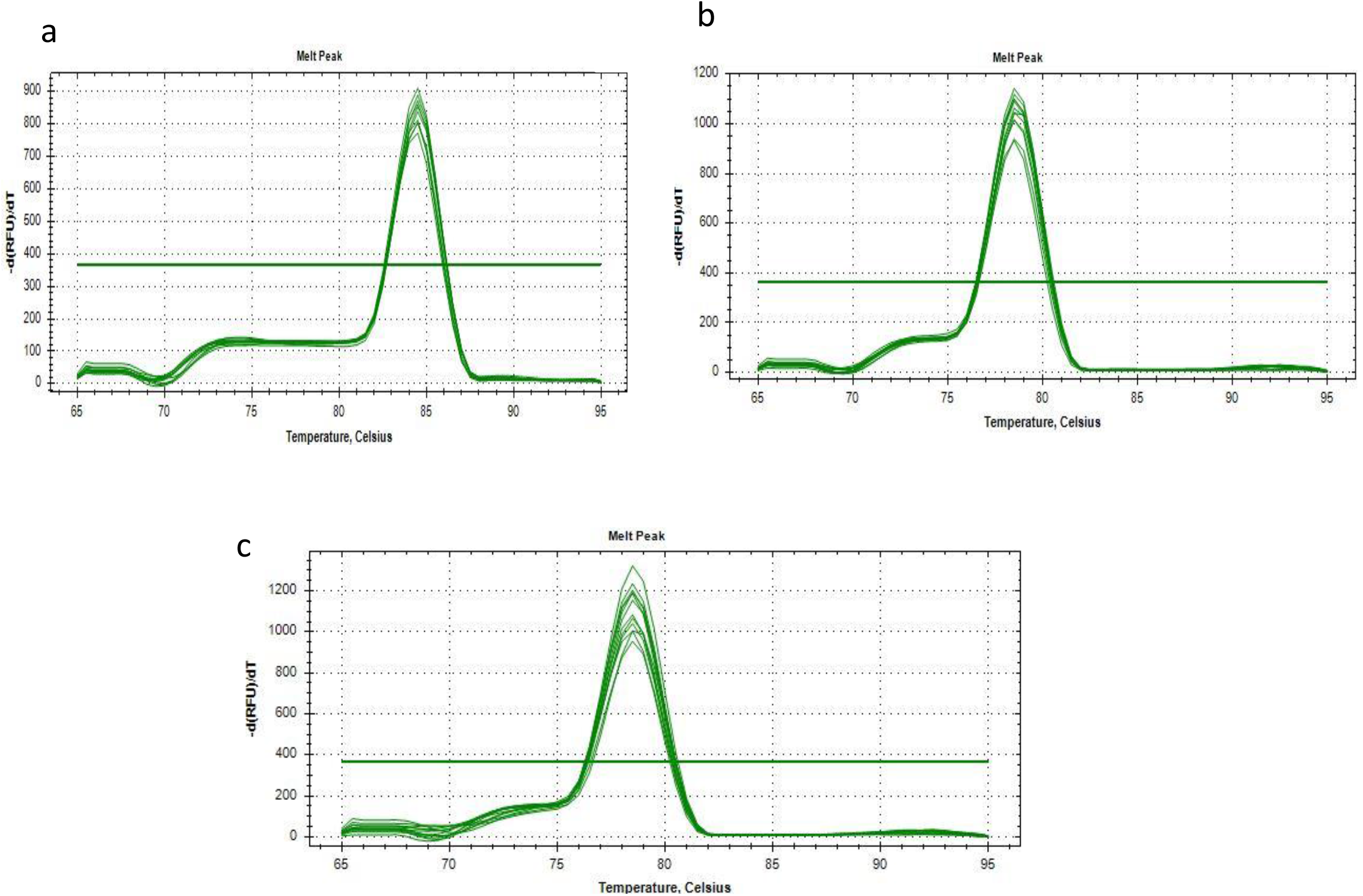
The Localization and design of the primers used. The nine nucleotides deleted in S gene and in N gene of Omicron variant are marked in bold.

### 2.4. Genome sequencing and lineage analysis

Ten samples displaying the Omicron variant profile by RT-PCR analysis were selected for genome sequencing. Samples were sequenced using the Nanopore MinION methodology and consensus sequences were generated using the bioinformatics SOP (https://artic.network/ncov-2019/ncov2019-bioinformatics-sop.html) in Nanopolish mode. Sequences obtained were deposited in GISAID database under the accession numbers EPI_ISL_7651286 to EPI_ISL_7651289, EPI_ISL_11327269, EPI_ISL_11327268, EPI_ISL_11327263, EPI_ISL_11327262, EPI_ISL_11327252 and EPI_ISL_11327251, were collected from 03 to 06 December 2021.

## 3. Results

Surveillance data, in the Laboratory of Molecular Biology and Cancer Immunology at the Lebanese University, revealed, the re-emergence of SGTF profile with a traveler coming from Abidjan on December 3, 2021. This sample was identified as Omicron BA.1 by whole genome sequencing. Since then, the number and the proportion of samples with SGTF profile were steadily increasing and have reached, as of January 10, approximately 95% of positive cases among passengers coming to Lebanon. This dramatic increase in SGTF percentage, suspected as Omicron BA.1 variant, urge the need for a rapid and accurate diagnostic tool to detect and track this variant. In this study, we developed a SYBR Green-based RT-qPCR assay for the detection of Omicron BA.1 variant. Our design was based on the differences in gene sequence from the original SARS-CoV-2 sequence (Fig. 1, Table 1).

The Omicron BA.1 contains several specific mutations that could potentially serve as a good tool for timely detection. These include a nine nucleotide deletions in the S gene Δ143-145, and other nine nucleotide deletions in the N gene Δ31-33. Accordingly, two groups of primers were designated. The first group detect the two mutations (spike del 143-145, and N del 31-33) was designed to detect these two mutations whereas, the second group (spike WT 143-145 and N WT 31-33) was designed to detect variants not harboring these deletions. A last primer pair (spike control) was designed from within the S gene in a region common to all variants including Omicron, and was used as a control for cDNA synthesis and to ensure RNA integrity.

### Calibration curve and limit of detection

In order to validate their accuracy and efficacy, the newly designed primer set underwent optimization by testing them on a serial 10-fold dilution of cDNA template as follows: 50, 5, 0.5, 0.05, 0.005, 0.0005 and 0.00005 ng/μl. All diluted samples were tested in triplicates by the gold standard TaqMan RT-qPCR (TaqPath kit) and by our SYBR Green-based RT-qPCR protocol (Table 2).

**Table 2.** Determination of the sensitivity by 7 serials 10-fold dilutions. Diluted samples were tested by our SYBR Green-based protocol and by TaqMan RT-qPCR (TaqPath kit).

|  | **CT values (SYBR Green)** |  |  | **Ct values**<br>**(Taq Path kit, S**<br>**probe)** |
| --- | --- | --- | --- | --- |
| **Concentration of DNA**<br>**Dilution Factor 1/10** | **Spike del 143-145** | **N del 31-33** | **S control** |  |
| **50 ng/μl** | 19.51 | 19.43 | 19.07 | 17.73 |
| **5 ng/μl** | 21.89 | 21.59 | 21.42 | 20.79 |
| **0.5 ng/μl** | 24.99 | 25 | 24.65 | 24.05 |
| **0.05 ng/μl** | 28.58 | 28.69 | 28.34 | 27.89 |
| **0.005 ng/ul** | 30.94 | 32.1 | 33.52 | 31.61 |
| **0.0005ng/ul** | 35.56 | 35.4 | 34.32 | 34.05 |
| **0.00005ng/ul** | ND | ND | ND | ND |

ND: Not Detected.

A calibration curve was generated for all the primer sets listed in table 2, using the serial dilution results. Linear regression performed for these primers sets demonstrated strong correlation (Fig. 2). A limit of detection (LOD) was determined for each primer set and identified as 0.0005 ng/μL for the two sets (Table 2, Fig. 2), or with maximal Ct of 34.05 or 35.5 obtained by TaqMan and SYBR Green RT-PCR respectively, resulting in a highly sensitive detection.

**Fig 2.**
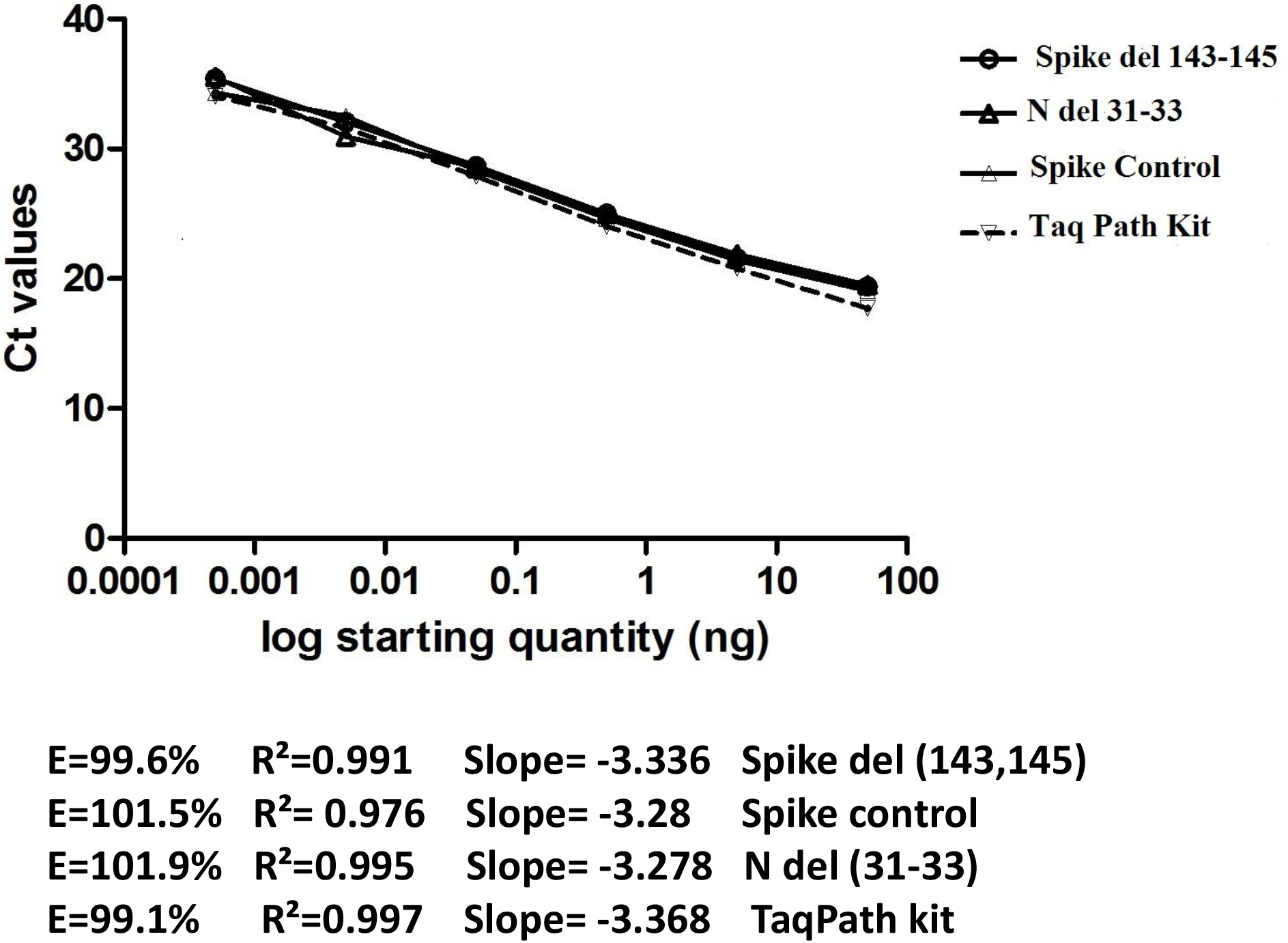
Calibration curves of SYBR Green-based qPCR primers Spike del 143-145, N del 31-33, S control and TaqPath kit. Serially 10-fold diluted cDNA containing the mutations S Δ143-145 and N Δ31-33 were amplified and analyzed. a) The threshold cycle (Ct) mean values were plotted against the Log starting quantity (ng/μl). slope, R2 and Eff were determined. Each dilution was assayed in triplicate.

Moreover, the specificity of our primers was investigated by three different ways: i) using 100 clinical negative samples for SARS-CoV-2 by TaqPath kit; they did not manifest a signal when tested with such negative controls. ii) using *in silico* prediction analyses as described in the Material and Methods section, and iii) by WGS of 10 samples.

To avoid any false-positive signals resulting from nonspecific products or primer-dimers, a melting curve analysis was included at the end of each PCR assay to determine the specificity and efficiency of each RT-qPCR reaction. Accuracy of our primers to amplify single amplicon is represented as single peak in melting curves (Fig. 3).

**Fig 3.**
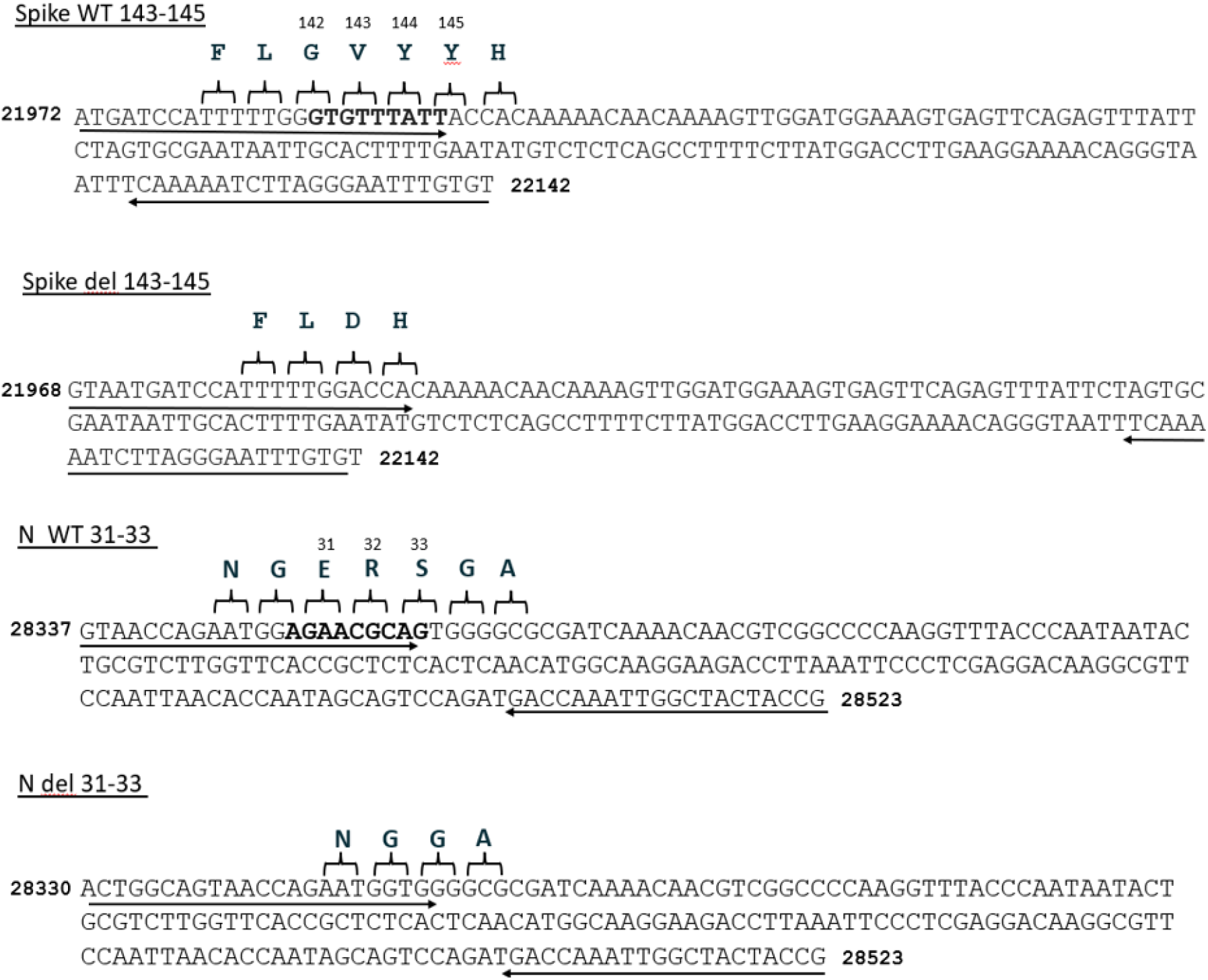
The melting curves for the products amplified with the SYBR Green-based qPCR protocol in S-gene target failure samples using the primers: a) Spike del 143-145; b) N del 31-33 and c) Spike control, reveal specific melt peak for each primer set.

### Clinical evaluation of Spike del 143-145 and N del 31-33 primer sets

After full optimization of the newly designed primer sets and the verification of their sensitivity and specificity in our qPCR assay, they were employed on 120 clinical samples positive for SARS-CoV-2, collected between 03 and 22 December 2021, and which have displayed a Ct < 30 with an SGTF profile, as revealed by the TaqPath kit. Our results showed that the spike control-gene amplicons were detected in all samples, providing evidence of cDNA synthesis and sample integrity preservation (Online Resource 1). Importantly, the S del 143-145-gene amplicons were detected in all the 120 samples, indicating the presence of the S Δ143-145 deletion and obviously the absence of the amino acids 143, 144 and 145 in the spike protein of these samples (Online Resource 1). Similar results were obtained for the N del 31-33-gene, amplicons were also detected in all the 120 samples indicating the presence of the N Δ31-33 deletion and the absence of the amino acids 31, 32 and 33 in the nucleocapsid protein of these samples. The use of S WT 143-145 and N WT 31-33 primers did not result in amplified product in any of the samples, thereby indicating that the S Δ143-145 and N Δ 31-33 deletions are located within the genomic region targeted by these two primer sets.

The amplification results obtained with spike del 143-145 and N del 31-33 primers were fully concordant with S WT 143-145 and N WT 31-33 primers, 100% of the SGTF samples harboring both S Δ143-145 and the N Δ 31-33 deletions (Online Resource 1).

Furthermore, ten of the S-negative samples were sequenced by WGS at the Microbial Pathogenomics Laboratory of the Lebanese American University (LAU) and sequences obtained were deposited in GISAID database under the accession number EPI_ISL_7651286 to EPI_ISL_7651289, EPI_ISL_11327269, EPI_ISL_11327268, EPI_ISL_11327263, EPI_ISL_11327262, EPI_ISL_11327252 and EPI_ISL_11327251. Sequence data analysis revealed that the ten samples belonged to the Omicron variant, while analyses of more sequences are in progress.

## 4. Conclusion

The protocol we described herein is faster, simpler and more cost-effective than genome sequencing-based method. The uniqueness of the targeted mutations (S Δ143-145 and N Δ31-33) renders this protocol highly accurate and amply adequate to the early identification of the suspected samples as Omicron BA.1 when genome sequencing is unavailable. In conclusion, this mutation-specific PCR assay would allow any laboratory having the ability to conduct PCR assays to rapidly and reliably screen for Omicron BA.1 among all SARS-CoV-2 positive samples with SGTF profile, in order to enhance surveillance capacity to identify cases and support decision making for interrupting transmission.

## Supporting information

Online Resource 1

## Data Availability

All data produced in the present study are available upon reasonable request to the authors

## Author contributions

FAS and KH Conceived and designed the experiments. RM, AK, NH, NB, ZAK, AH, NK, HK, LR, NS, FAN, ZG, NEZ and EH Performed the molecular biology assays. KH and FAS Analyzed and interpreted the data. The first draft of the manuscript was written by KH, FAS and NH. ZAK, AH and NK commented on the first draft of the manuscript. All authors read and approved the final manuscript.

## Availability of data and materials

We declared that materials described in the manuscript, including all relevant raw data, will be freely available to any scientist wishing to use them for non-commercial purposes, without breaching participant confidentiality.

## Ethics approval and consent to participate

All presented cases or their legal guardian provided consent to data collection and use according to institutional guidelines.

## Consent for publication

All presented cases or their legal guardian provided consent to publish according to institutional guidelines.

