## Supplementary material for "Rapid detection of SARS-CoV-2 B.1.1.529 (Omicron BA.1) variant by SYBR Green□based RT□qPCR": Online Resource 1

**Corresponding author**:

^$^ Co-corresponding authors

**Online Resource 1.** The Ct values from our SYBR Green-Based assay protocol.

|  | **Ct Values (TaqPath kit)** | **Ct values (SYBR Green- Based)** | | | | |
| --- | --- | --- | --- | --- | --- | --- |
| **samples** | **ORF1ab probe** | **N WT 31-33** | **N del 31-33** | **S WT 143-145** | **S del 143-145** | **spike control** |
| 1 | 21.6 | ND | 24.39 | ND | 23.46 | 25.6 |
| 2 | 15.08 | ND | 17.21 | ND | 16.45 | 16.93 |
| 3 | 18.8 | ND | 19.57 | ND | 19.21 | 18.65 |
| 4 | 25.7 | ND | 26.87 | ND | 25.57 | 25.87 |
| 5 | 21.58 | ND | 23.68 | ND | 23.2 | 22.46 |
| 6 | 17.2 | ND | 18.49 | ND | 18.06 | 18.31 |
| 7 | 17.4 | ND | 17.15 | ND | 17.11 | 17.69 |
| 8 | 25.27 | ND | 27.08 | ND | 25.39 | 25.78 |
| 9 | 24.003 | ND | 26.9 | ND | 25.27 | 25.12 |
| 10 | 18.5 | ND | 22.31 | ND | 20.31 | 21.28 |
| 11 | 18.46 | ND | 19.96 | ND | 18.69 | 20.16 |
| 12 | 19.94 | ND | 20.9 | ND | 19.47 | 20.29 |
| 13 | 17.65 | ND | 18.53 | ND | 18.06 | 19.17 |
| 14 | 21.6 | ND | 22.02 | ND | 21.7 | 21.91 |
| 15 | 23.32 | ND | 23.73 | ND | 23.38 | 24.19 |
| 16 | 23.54 | ND | 24.82 | ND | 24.54 | 25.32 |
| 17 | 18.21 | ND | 19.54 | ND | 18.54 | 19.01 |
| 18 | 18.03 | ND | 17.49 | ND | 17.08 | 17.88 |
| 19 | 22.69 | ND | 21.17 | ND | 21.64 | 22.05 |
| 20 | 23.59 | ND | 27.29 | ND | 26.78 | 26.84 |
| 21 | 16.6 | ND | 18.71 | ND | 19.16 | 19.27 |
| 22 | 20.5 | ND | 22.2 | ND | 22.46 | 23.02 |
| 23 | 26.69 | ND | 28.61 | ND | 28.92 | 29.02 |
| 24 | 20.6 | ND | 22.45 | ND | 22.8 | 23.34 |
| 25 | 16.8 | ND | 20.55 | ND | 20.12 | 19.78 |
| 26 | 18.69 | ND | 21.18 | ND | 21.03 | 20.28 |
| 27 | 23.02 | ND | 24.9 | ND | 24.93 | 24.81 |
| 28 | 25.64 | ND | 27.85 | ND | 27.49 | 27.53 |
| 29 | 20.8 | ND | 22.26 | ND | 22.85 | 23.42 |
| 30 | 24.01 | ND | 27.5 | ND | 27.43 | 27.44 |
| 31 | 25.28 | ND | 27.53 | ND | 27.76 | 28.4 |
| 32 | 22.8 | ND | 28.84 | ND | 25.11 | 25.07 |
| 33 | 21.69 | ND | 24.57 | ND | 24.24 | 24.35 |
| 34 | 24.71 | ND | 26.18 | ND | 27.08 | 27.03 |
| 35 | 30.07 | ND | 34.33 | ND | 33.03 | 32.88 |
| 36 | 22.12 | ND | 25.4 | ND | 25.34 | 25.06 |
| 37 | 26.14 | ND | 27.79 | ND | 29.6 | 29.6 |
| 38 | 21.1 | ND | 24.56 | ND | 25.14 | 25.1 |
| 39 | 25.13 | ND | 30.45 | ND | 30.01 | 30.23 |
| 40 | 19.6 | ND | 25 | ND | 25.16 | 25.21 |
| 41 | 20.24 | ND | 24.94 | ND | 26.67 | 27.22 |
| 42 | 25.7 | ND | 31.34 | ND | 33.21 | 33.79 |
| 43 | 25.9 | ND | 31.15 | ND | 31.63 | 33.04 |
| 44 | 22.82 | ND | 25.16 | ND | 25.78 | 25.59 |
| 45 | 28.58 | ND | 31.83 | ND | 31.77 | 31.45 |
| 46 | 16.52 | ND | 21.17 | ND | 22.59 | 22.7 |
| 47 | 21.25 | ND | 25.69 | ND | 27.5 | 28.02 |
| 48 | 28.3 | ND | 29.47 | ND | 30.23 | 30.28 |
| 49 | 18.96 | ND | 22.28 | ND | 24.34 | 24.52 |
| 50 | 26.71 | ND | 28.77 | ND | 29.97 | 30.52 |
| 51 | 29.17 | ND | 29.8 | ND | 32.52 | 33.46 |
| 52 | 20.9 | ND | 23.39 | ND | 26.05 | 26.4 |
| 53 | 18.86 | ND | 21.17 | ND | 23.03 | 23.07 |
| 54 | 18.03 | ND | 19.8 | ND | 22.54 | 24.2 |
| 55 | 20.9 | ND | 22.42 | ND | 23.67 | 24.09 |
| 56 | 28.3 | ND | 31.92 | ND | 32.53 | 32.92 |
| 57 | 17.13 | ND | 20.52 | ND | 22.58 | 22.52 |
| 58 | 20.5 | ND | 25.24 | ND | 26.1 | 26.12 |
| 59 | 25.3 | ND | 28.09 | ND | 30.16 | 32.56 |
| 60 | 20.09 | ND | 22.18 | ND | 24.06 | 24.79 |
| 61 | 29.76 | ND | 32.51 | ND | 34.66 | 33.41 |
| 62 | 14.4 | ND | 18.25 | ND | 18.63 | 19.14 |
| 63 | 21.11 | ND | 22.47 | ND | 25.2 | 25.36 |
| 64 | 19.95 | ND | 21.2 | ND | 24.37 | 25.55 |
| 65 | 20.26 | ND | 22.43 | ND | 23.76 | 23.94 |
| 66 | 21.8 | ND | 24.21 | ND | 25.34 | 25.21 |
| 67 | 28.6 | ND | 32.83 | ND | 34.31 | 33.5 |
| 68 | 20.72 | ND | 23.17 | ND | 22.99 | 22.66 |
| 69 | 27.02 | ND | 31.83 | ND | 31.21 | 32.72 |
| 70 | 29.18 | ND | 32.98 | ND | 33.09 | 33.5 |
| 71 | 24.42 | ND | 27.73 | ND | 30.12 | 30.1 |
| 72 | 18.74 | ND | 21.71 | ND | 24.03 | 25.16 |
| 73 | 16.85 | ND | 20.53 | ND | 21.9 | 21.6 |
| 74 | 17.56 | ND | 22.84 | ND | 23.09 | 23.37 |
| 75 | 22.98 | ND | 24.86 | ND | 25.52 | 25.71 |
| 76 | 20.7 | ND | 24.51 | ND | 24.53 | 20.02 |
| 77 | 20.33 | ND | 21.58 | ND | 23.34 | 23.9 |
| 78 | 24.51 | ND | 27.13 | ND | 28.95 | 28.46 |
| 79 | 23.57 | ND | 26.66 | ND | 27.49 | 27.27 |
| 80 | 20.2 | ND | 22.31 | ND | 24.11 | 24.61 |
| 81 | 29.4 | ND | 31.25 | ND | 33.16 | 32.5 |
| 82 | 19.9 | ND | 21.37 | ND | 22.46 | 23.05 |
| 83 | 20.3 | ND | 23.29 | ND | 24.88 | 24.3 |
| 84 | 26.68 | ND | 28.65 | ND | 30.36 | 32.17 |
| 85 | 22.98 | ND | 25.91 | ND | 26.98 | 26.74 |
| 86 | 16.97 | ND | 20.02 | ND | 19.65 | 20.2 |
| 87 | 18.5 | ND | 20.65 | ND | 21.16 | 21.92 |
| 88 | 30.1 | ND | 33.67 | ND | 32.36 | 33.57 |
| 89 | 23.7 | ND | 27.07 | ND | 26.74 | 26.57 |
| 90 | 21.24 | ND | 23.86 | ND | 25.41 | 25.57 |
| 91 | 26.77 | ND | 27.94 | ND | 29.18 | 29.74 |
| 92 | 24.18 | ND | 25.58 | ND | 26.71 | 26.42 |
| 93 | 20.77 | ND | 23.13 | ND | 24.03 | 23.75 |
| 94 | 26.92 | ND | 28.26 | ND | 29.32 | 28.86 |
| 95 | 24.2 | ND | 25.25 | ND | 27.31 | 27.04 |
| 96 | 20.85 | ND | 20.26 | ND | 22.51 | 22.18 |
| 97 | 28.51 | ND | 29.89 | ND | 31.26 | 21.34 |
| 98 | 29.81 | ND | 30.72 | ND | 32.9 | 32.37 |
| 99 | 29.03 | ND | 31.17 | ND | 32.26 | 31.93 |
| 100 | 23.85 | ND | 20.58 | ND | 22.66 | 22.27 |
| 101 | 28.2 | ND | 29.43 | ND | 30.32 | 31.23 |
| 102 | 25.14 | ND | 27.58 | ND | 27.9 | 27.6 |
| 103 | 17.42 | ND | 17.15 | ND | 18.32 | 18.31 |
| 104 | 30.69 | ND | 32.37 | ND | 33.17 | 33.43 |
| 105 | 26.99 | ND | 27.26 | ND | 27.64 | 27.54 |
| 106 | 30.32 | ND | 31.1 | ND | 31.87 | 34.65 |
| 107 | 26.93 | ND | 26.19 | ND | 28.74 | 28.82 |
| 108 | 22.6 | ND | 23.73 | ND | 25.23 | 25.5 |
| 109 | 23.47 | ND | 26.58 | ND | 27.29 | 27.9 |
| 110 | 20.22 | ND | 21.79 | ND | 22 | 21.54 |
| 111 | 22.52 | ND | 23.76 | ND | 24.1 | 23.95 |
| 112 | 23.55 | ND | 24.33 | ND | 25.06 | 24.1 |
| 113 | 24.11 | ND | 28.28 | ND | 29.26 | 29.4 |
| 114 | 28.68 | ND | 28.23 | ND | 29.38 | 29.19 |
| 115 | 25.58 | ND | 25.67 | ND | 27.02 | 27.59 |
| 116 | 19.64 | ND | 23.74 | ND | 24.61 | 24.41 |
| 117 | 27.38 | ND | 28.3 | ND | 28.6 | 28.65 |
| 118 | 21.14 | ND | 20.47 | ND | 21.03 | 20.82 |
| 119 | 19.54 | ND | 20.53 | ND | 21.9 | 21.6 |
| 120 | 20.15 | ND | 22.84 | ND | 23.09 | 23.37 |

ND: Not Detected.
